# Inclusion bias affects common variant discovery and replication in a health-system linked biobank

**DOI:** 10.1101/2025.04.04.25325131

**Authors:** Aditya Pimplaskar, Junqiong Qiu, Sandra Lapinska, Veronica Tozzo, Jeffrey N. Chiang, Bogdan Pasaniuc, Loes M. Olde Loohuis

## Abstract

Electronic Health Records (EHR)-linked biobanks have emerged as promising tools for precision medicine, enabling the integration of clinical and molecular data for individual risk assessment. Association studies performed in biobank studies can connect common genetic variation to clinical phenotypes, such as through the use of polygenic scores (PGS), which are starting to have utility in aiding clinician decision making. However, while biobanks aggregate large amounts of data effectively for such studies, most employ various opt-in consent protocols, and, as a result, are expected to be subject to participation and recruitment biases. The extent to which biases affect genetic analyses in biobanks remains unstudied. In this study, we quantify bias and evaluate its impact on genetic analyses, using the UCLA ATLAS Community Health Initiative as a case study. Our analyses reveal that a wide array of factors, particularly socio-demographic characteristics and healthcare utilization patterns, influence participation, effectively differentiating biobank participants from the broader patient population (AUROC = 0.85, AUPRC = 0.82). Through weighting the sample using inverse probability weights derived from probabilities of enrollment, we replicated 54% more known GWAS variants than models that did not take bias into account (e.g. associations between variants in the *PPARG* gene and type 2 diabetes). We further show that PGS-Phenome wide associations are affected by the weighting scheme, and suggest associations corroborated by weighted analyses to be more robust. Our results highlight that genetic analyses within biobanks should account for inclusion biases, and suggest inverse probability weighting as a potential approach.

## Introduction

Electronic Health Records (EHR)-linked biobanks from health systems are becoming increasingly common and powerful^1,2^, enabling the discovery of phenotypic and genetic associations with clinical features, offering promise for precision medicine^3,4,5,6^. These biobanks facilitate personalized risk assessments, leveraging clinical risk factors learned from health records and molecular data to compute polygenic scores (PGS) as estimates of genetic risk. These PGS can be used to distinguish cases from controls^7–9^, as well as for subtyping within case groups^10–13^ and informing interventions and treatment decisions^14,15^. Like most large research resources, biobanks typically employ opt-in consent protocols and use a variety of recruitment strategies. These recruitment and consent protocols can induce biases in biobank cohorts which may impact genetic analyses^16^ and the clinical utility of risk assessments^17^. Several studies have compared participants of population-based cohort studies to the background population using publicly available census data. In the UK Biobank, these studies identified a “healthy participant” bias^18,19^, and demonstrated its effect on the prediction of clinical outcomes, association of lifestyle and demographic factors to health^20,21^, and estimates of heritability and genetic correlation between traits^22^.

In contrast to these population-based studies, EHR-linked biobanks recruit participants through their interaction with a healthcare system; as a result, disease severity and greater healthcare utilization^23^, along with other demographic factors, may differentiate enrolled individuals from the broader population. The degree of inclusion biases in EHR-linked biorepositories, and the downstream impact on genetic analyses of these biases remain unexplored.

In this study, we use data from the UCLA ATLAS Community Health Initiative’s biobank (ATLAS), which is part of the greater UCLA Health system serving the Los Angeles area. The UCLA EHR consists of health record data for over 4 million patients, 104,516 of whom have enrolled in the ATLAS initiative^24,25^. The health records span a variety of care settings and domains. ATLAS began recruitment in 2016, initially with opt-in protocols in operative and blood testing settings, which were later extended to widespread recruitment efforts using digital messaging through patient portals^26^.

In this study, we propose a framework to assess inclusion biases in biobank studies, test and mitigate their impact on genetic investigations. We performed univariate association analyses between clinical, demographic, and healthcare utilization features and ATLAS enrollment. Using these features, we developed classification models of enrollment to distinguish the ATLAS-enrolled group from the background population. Using the probabilities derived from these models, we generated inverse-probability weights. With these weights, we then tested the effects of bias on discovery tasks, including common variant replication and PGS-phenome-wide associations (PGS-PheWAS). Our results show that models trained on demographics, healthcare utilization, and diagnostic data can distinguish biobank participants from the background population (AUROC=0.85), as hypothesized. By incorporating inverse probability weights, we showed that accounting for enrollment bias increased the replication rate of known GWAS associations in ATLAS by 54% and altered the results of PGS-PheWAS scans. Our results suggest that using weighted models for bias adjustment to corroborate discovery in common variant studies may lead to more robust associations within the study context.

## Results

### Cohort characteristics

From the UCLA Electronic Health Records (EHR), encompassing a total sample of n=4,310,043 patients in the UCLA Health system, we retained n=3,734,854 after filtering out individuals whose records ended prior to the start of ATLAS recruitment and with missing demographic information. We further restricted the sample to individuals aged 18-90 in the span of ATLAS’ recruitment period, in accordance with the enrollment consent protocol for the biobank, with at least one visit in which a ICD-10 diagnostic code was recorded. This resulted in a sample of n=1,568,927 patients, of whom n=104,516 (6.7%) individuals consented to participate in the UCLA ATLAS study by the end of 2023. Participants who provided consent for enrollment in the ATLAS sample allowed for residual tissue samples from future clinical visits to be saved for genotyping and further research use; of the 104,516 ATLAS participants, 54,770 had samples collected and quality-controlled genotypes available at the time of analysis^25^.

For the included sample, we extracted patient sex, age at latest visit, whether or not the patient received primary care at UCLA, frequency of clinical contacts, “Social Vulnerability Index” (SVI)^27^, “Barriers to Accessing Services” (BAS)^28^ indicator, self-reported race and ethnicity (encoded as Ethnoracial category), insurance categories associated with clinical visits (grouped into Federal, State, Private, or None), smoking status, and medical history. Features were extracted prior to the date of enrollment for all ATLAS-enrolled patients and prior to the end of 2023 for all unenrolled.

### Univariate associations indicate significant enrollment-specific covariates

We compared the distributions and assessed the impact of each of our demographic and clinical features by using univariate logistic regression models with the enrollment outcome, and identified various signatures of the enrolled patient population (Figure 1, Supplementary Taable 1). For example, UCLA Health patients who enroll in ATLAS tend to self-report as White and Non-Hispanic with 57.3% of enrolled individuals self-reporting as White, compared to 43% in the unenrolled sample. (OR_Black/African-American = 0.604, 95% CI: [0.58-0.62], OR_Hispanic/Latino = 0.76, 95% CI: [0.74-0.77]). ATLAS patients are likely to receive primary care at UCLA (70.2%), compared to 21.8% of the unenrolled sample (OR=8.44, 95% CI: [8.33-8.56]); this is in line with ATLAS participants being more likely (88.4% versus 50.2% in the unenrolled sample) to receive a Z0 ICD-10 diagnosis (OR = 7.57, 95% CI: [7.43-7.72]) labeled “Encounter for general examination without complaint, suspected or reported diagnosis”.

**Figure 1:**
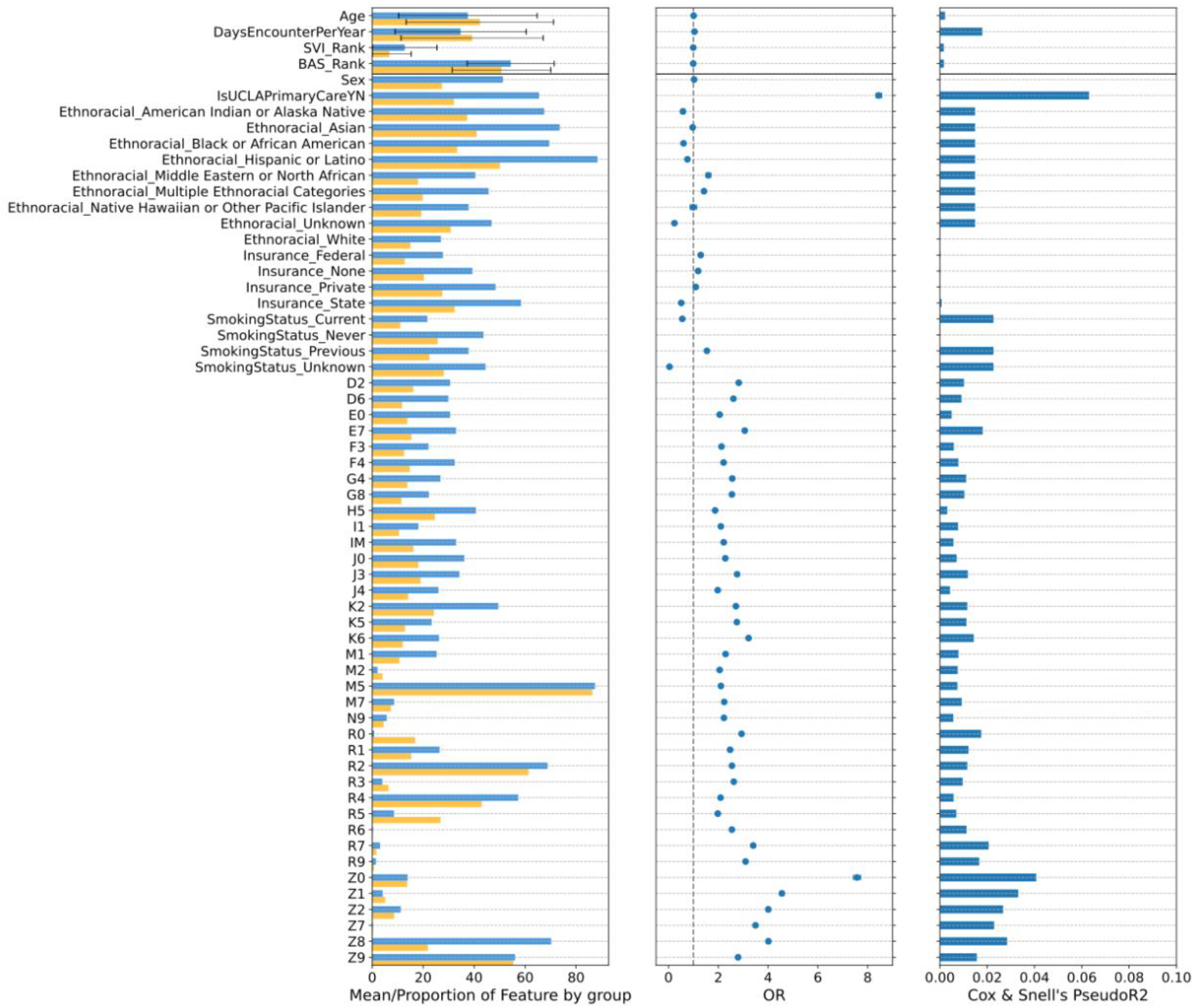
Cohort characteristics of the UCLA health sample and ATLAS subsample. (Left) Feature level distributions stratified by ATLAS enrollment (yellow = not enrolled in ATLAS, blue = enrolled in ATLAS). For quantitative variables, feature means are depicted, and for categorical features, proportion of individuals in both groups in the feature group are depicted. For diagnostic chapters, any diagnoses in the chapter is sufficient. (Middle/right) Results from univariate associations with ATLAS enrollment Odds ratio (middle) and Cox & Snell’s pseudo-R2 (right).

Previous smokers were overrepresented in the ATLAS enrolled sample with a prevalence of 26.4% compared to 15.3% in the unenrolled group (OR=1.54, 95% CI = [1.52-1.57]), but current smokers were underrepresented in ATLAS with a prevalence of 3.97% compared to 6.4% in the unenrolled sample (OR = 0.55, 95% CI = [0.53 - 0.57]). While most patients use private health insurance at UCLA, ATLAS participants were found to be less likely to use state insurance (2.08%) compared to 3.98% in the unenrolled group (OR=0.51, 95% CI: [0.49-0.53]). ATLAS participants further showed a higher diagnostic burden, though these trends in diagnostic burden were not confined to any particular diagnostic category. This reflects a more global increase in diagnostic burden in ATLAS enrolled individuals compared to the background UCLA Health population (Supplementary Figure 1), which was also reflected in the overall increased number of days with visits per year of ATLAS participants (ATLAS Enrolled: 12.8, Unenrolled: 6.7, OR=1.044, 95% CI: [1.043-1.045]).

We further performed sensitivity analyses to evaluate how feature level differences between ATLAS participants and non-participants changed over time, and were affected by healthcare utilization. Since the ATLAS biobank’s inception, various strategic changes have been implemented in the recruitment protocol; these include transitions from in-person recruitment in inpatient clinics to dissemination of universal consent forms via online patient portals as well as modifications due to COVID-19 restrictions. Across various combinations of healthcare utilization (thresholded by number of visits) and ATLAS recruitment timeframes, we evaluated the direction and magnitude of effect of each feature, and found that feature effects predominantly remain consistent (Supplementary Figure 2)

### Multivariate random-forest models to estimate enrollment probability

In order to test our ability to detect enrollment from all available features, we trained random forest models, allowing us to account for feature effects jointly to retrieve reliable enrollment probabilities. With a 10-fold cross-validation AUC of 0.85 (AUPRC = 0.82) (Supplementary Figure 3), the random forest model distinguished the ATLAS-enrolled sample from other UCLA Health patients. While the majority of ATLAS-enrolled individuals have markedly higher predicted probabilities of enrollment, the distributions of predicted probability for ATLAS and non-ATLAS participants overlap, with some ATLAS participants having been assigned low probabilities of enrollment by the random forest, and vice versa (Figure 2). We assessed feature importances with Shapley values, quantifying feature contributions on model predictions while accounting for other model features. Highly important features in the model were consistent with findings from univariate associations and included receiving primary care at UCLA Health, the frequency of healthcare utilization, while also implicating several ICD-10 codes of abnormalities in imaging, blood testing, and metabolic disorders (Figure 2).

**Figure 2:**
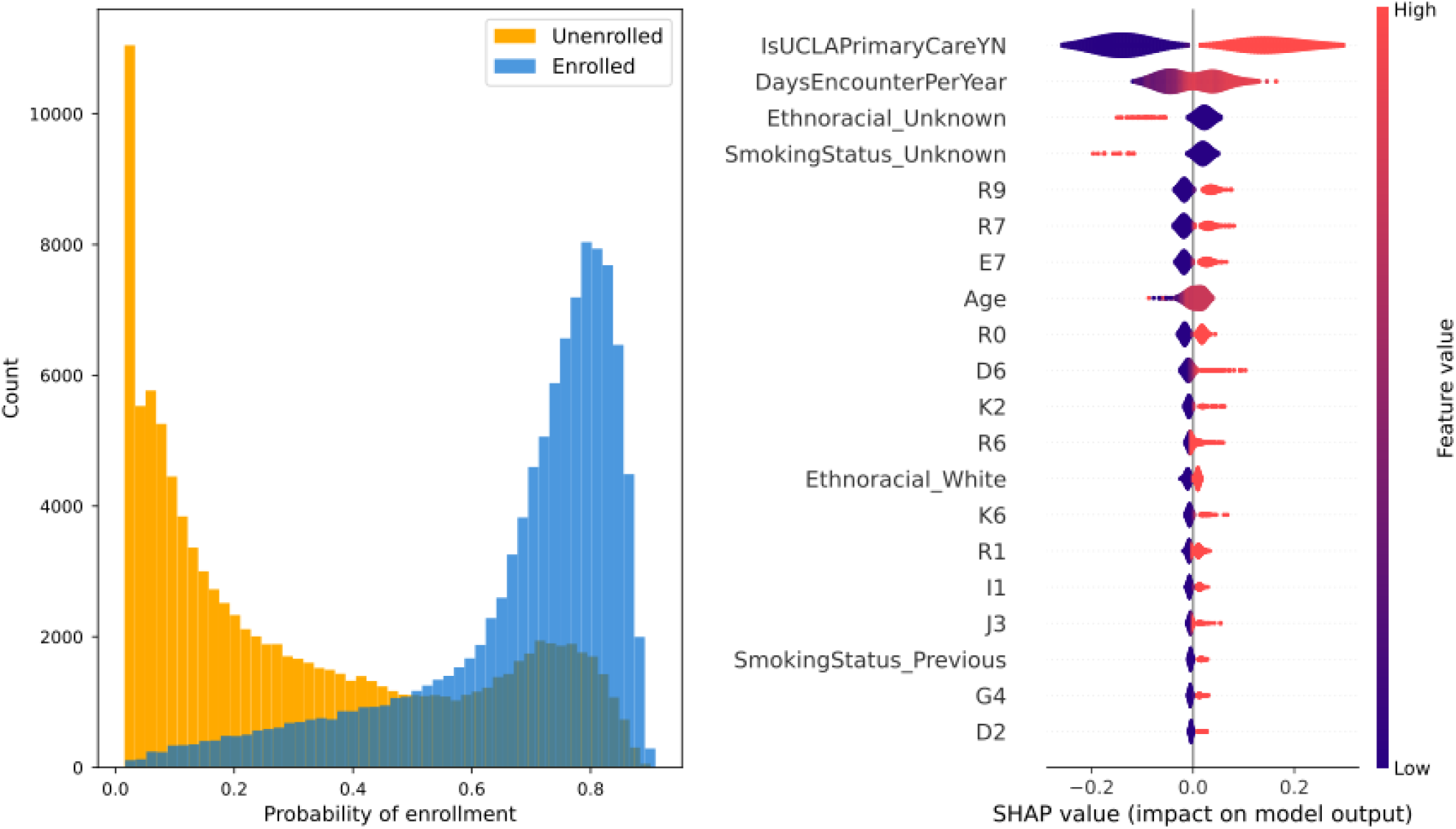
Probability distributions stratified by enrollment and feature characteristics from a multivariate random forest model classifier of ATLAS enrollment. (Left) Predicted probability distributions stratified by true ATLAS enrollment status (yellow = not enrolled in ATLAS, blue = enrolled in ATLAS) show strong separation between the classes. (Right) Beeswarm plot of Shapley values on a subset of 100 individuals reveal healthcare utilization patterns and select ICD-10 diagnoses as important predictors of ATLAS enrollment.

Through recursive feature elimination (RFE), we constructed models that, based on reduced sets of predictors (with 5, 10, and 15 features), still identified ATLAS enrolled individuals with high accuracy (Supplementary Figure 3, Supplementary Table 2). A 5-feature model using age at latest visit, primary care status, frequency of healthcare encounters per year, and indices for social vulnerability (SVI) and barriers to accessing service (BAS) as predictors distinguished ATLAS-enrolled from unenrolled individuals with a performance very similar to the full model (AUC=0.82, AUPRC = 0.78). This model was used downstream for generating inverse-probability weights.

Since receiving primary care at UCLA was the strongest predictor in our model, we evaluated classification models including only the subset of primary care patients at UCLA. While overall classification performance was reduced (AUC of 0.69, AUPRC = 0.68), performance was moderate, and univariate associations were in line with the larger model (See Supplementary Table 3, Supplementary Figure 4).

### Weights and readjusting univariate associations

Predicted probabilities of enrollment were recovered from the 5-feature reduced random-forest model, after which a transformation and normalization was performed to generate inverse-probability weights (Supplementary Figure 5). The weights were then used to create a weighted ATLAS sample more representative of the background UCLA Health population. We tested whether adjusting by re-weighting effectively removed the previously observed associations between univariate features and enrollment and observed that prior associations were no longer significant (Supplementary Figure 5).

### Replicability of known GWAS variants is improved under weighting scheme

Next, we tested whether accounting for the effect of inclusion biases impacts the ability to replicate known common trait-variant associations in ATLAS. To do so, we used the phenotype-genotype reference map from *pgrm*^*29*^, which catalogs a curated set of variant-phecode associations, and compared replications of these known associations in unweighted and weighted settings. Out of the 5411 SNP-phecode associations in *pgrm*, 1879 (34.7%) met filtering criteria based on data availability in ATLAS and were tested for association (Supplementary Table 4). These associations included 27 phecodes, with the majority coming from phecodes 250.2 (type 2 diabetes), 495 (asthma), and 427.21 (atrial fibrillation). We considered an association replicated if it was significantly associated with p < 0.05 and effect directionality was consistent with *pgrm*.

Replicated associations spanned 24 phecodes (breakdown in Supplementary Figure 6). Under both unweighted and weighted settings, 389 (20.7%) associations were replicated. We observed 217 (11.5%) associations that were replicated only in the unweighted analyses, while 335 (17.8%) associations were replicated only in the weighted setting (an increase of 54%, p= 3.49e-8). When comparing all 724 weighted associations to the 606 unweighted associations (including those identified in both settings), the relative increase was 19.5% (p= 3.29e-5). The associations that replicated in both weighted and unweighted settings, 255 (67%) were more strongly associated under the weighted scheme (Table 1).

**Table 1:**
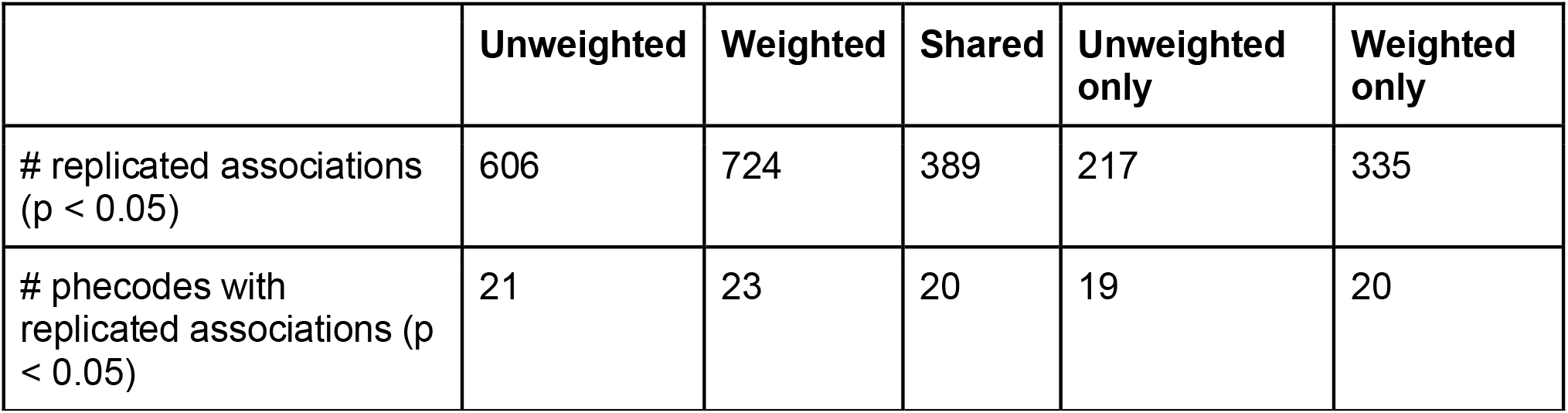
Counts and proportions of replicated associations from pgrm in the ATLAS sample. Associations were considered replicated at a significance level of p < 0.05 and when direction of effect was consistent with pgrm.

This trend of greater replication in the weighted setting is robust (Supplementary Figure 7), and even stronger, subject to the application of Bonferroni correction (threshold of p < 0.05/1879): 61 (3.2%) associations were replicated under both weighting schemes, 33 (1.8%) only in unweighted setting, and 114 (6.1%) only in the weighted setting, corresponding to an increase of 245% considering associations unique in the weighted/unweighted setting (Z-test p-value = 8.41e-12), versus 86.2% overall (Z-test p-value=2.07e-7).

Among the associations that replicated in the weighted model setting but failed to replicate in the unweighted setting were several well-established associations; for example, variants in the *PPARG* gene implicated in type 2 diabetes^30^ (intronic variant rs11709077, weighted p=2.57e-5, unweighted p=0.076; missense variant rs1801282 Weighted p=1.39e-4, Unweighted p=0.12) and in the *CELSR2-PRSC1-SORT1* gene cluster, associated with coronary atherosclerosis (rs7528419 lies in the 3’ UTR of *CELSR2*, Weighted p=3.47e-6, Unweighted p=0.10).

### Weighting scheme modifies observed phenotypic relationships in phenome-wide associations with polygenic risk

Biobanks are often used to study genetic architecture of phenotypes through PGS associations. To test the impact of inclusion bias on such analyses, we performed weighted and unweighted phenome-wide scans (Figure 3) associating a collection of 541 phecodes with the PGS for Major depressive disorder (MDD)^31^, and body mass index (BMI)^32^. These traits were selected due to their polygenic nature, availability of large GWAS, their frequent diagnosis/availability in the ATLAS sample, and their association with a myriad of comorbidities and associated health complications^33–37^. As expected, clinically relevant phecodes emerged as the most significantly associated with each respective PGS – ‘mood disorders’ for MDD and and ‘overweight, obesity, and other hyperalimentation’ for BMI. The effect of the weighting scheme was minimal on these associations which retained strong statistical significance and similar effect sizes (Supplementary Table 5).

**Figure 3:**
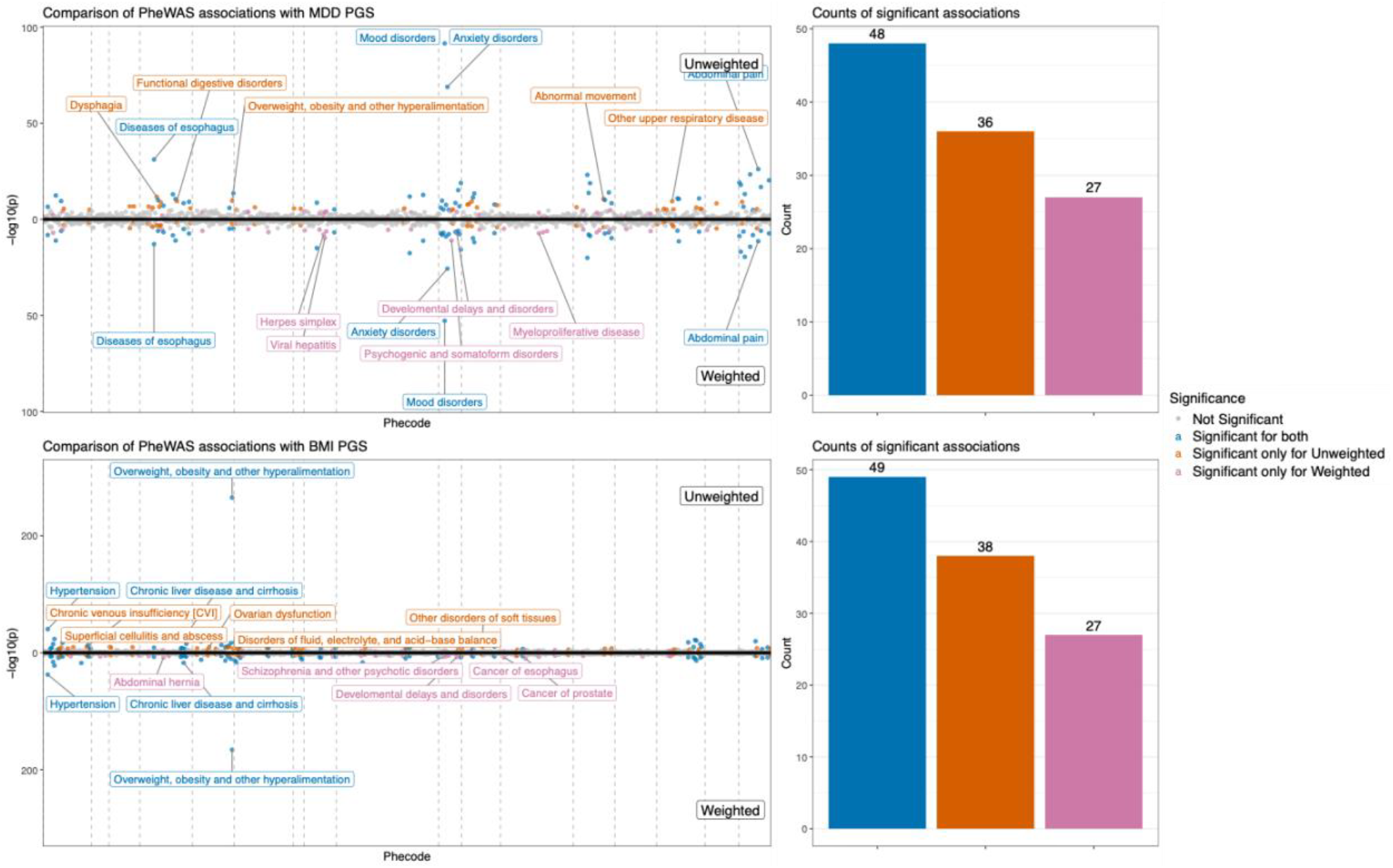
Comparison of weighting schemes in phenome-wide associations with PGS. Miami plots for PheWAS on PGS for MDD and BMI (top: unweighted, bottom: weighted) show shared and unique associations under the unweighted and weighted models.

For phenotypes that are closely associated with PGS traits, we observed an enrichment of associations that were significant both in unweighted and weighted settings (Supplementary Table 6). The ‘mental disorders’ phecode category was enriched for associations corroborated by the weighting scheme for the MDD PGS (Fisher’s exact test, p=0.0097). Further, in a subset of metabolic traits (including diabetes mellitus, nutritional deficiencies, and disorders of metabolism), associations with all of these traits were corroborated by the weighting scheme. We hypothesized that associations only detected in the unweighted setting may be more affected by features tagging the bias found in the biobank. To investigate this, we explicitly adjusted for the 5 features that comprise the reduced model from which weights were generated. From the 36 phecodes only associated with the MDD PGS in the unweighted setting alone, 36% are no longer significantly associated after explicitly accounting for the 5 covariates, compared to 14.3% of the 42 associations identified under both the weighted and unweighted settings. This distinction is even more stark in the BMI PGS setting, where 65.8% of the 38 associations significant under only the unweighted setting became non-significant compared to only 26.5% of the 49 associations that were significant under both the unweighted and weighted setting. Significance breakdown across phecode categories can be found in Supplementary Figure 8.

We further investigated whether the specific PheWAS associations only observed in the unweighted setting were attributable to any particular bias-inducing feature. To assess the effect of each of the 5 features from the reduced random forest model on the PGS-phecode associations found significant under the unweighted model setting, we parallelly performed an unweighted phenome-wide scan, appending the 5 features as additional covariates. We observed a heterogeneous pattern of association between each of the 5 features and the phecodes, indicating that the bias induced by these features on PGS-phecode associations varies by the phecode and genetic measure in question (Supplementary Figure 9). These findings are in line with the hypothesis that some of the associations found to be significant in an unweighted analysis may be attributable to features reflecting bias in the biobank, rather than true PGS-phecode association.

## Methods and Data

### Data querying & filtering

The UCLA Health Discovery Data Repository (DDR) is a de-identified data repository for UCLA Health system-wide data, consisting of electronic health records (EHR) with clinical information. This data spans 2 hospitals and 210 primary/specialty outpatient locations in the UCLA health system, serving around 5% of Los Angeles County. The UCLA ATLAS Precision Health Biobank is a data resource for research purposes, linking clinical data from the DDR to genomic data for a subset of UCLA Health patients. Patients who received healthcare through UCLA Health were invited to enroll in ATLAS through the universal consent process. Individuals considered in this study were recruited between 2016 through 2023, consistent with the latest available release of genetic data.

To generate our study cohort, we included data from all available healthcare locations recruiting a sample of 3,749,826 individuals who have had at least one encounter between 2016 and 2023 (the UCLA ATLAS recruitment period). We further filtered the sample by retaining only individuals aged at least 18 in 2023 and removed 14970 individuals with missing sex. At the time of analysis, 104,516 ATLAS participants had consented into the biobank with 54,770 having samples collected and quality-controlled genotypes; we further excluded all those who did not have phecode data available to generate a cohort of 48,664 samples with genetic and clinical data available for genetic analyses.

We further subset the cohort to those receiving primary care at UCLA (n=400,532) for use in sensitivity analysis, of whom 74,266 were enrolled in ATLAS and 326,266 were unenrolled.

### Feature extraction

We extracted features related to demographics, healthcare utilization, and clinical characteristics for both the ATLAS and reference UCLA Health populations. Demographic features include age at the latest visit, whether individuals received primary care at UCLA, indices for social vulnerability and access to care, ethnoracial category (a feature defined internally based on self-reported race and ethnicity to enhance clarity in categorization of self-reported race-related data), and smoking status. Healthcare utilization was encoded as the number of visits scaled by the length of an individual’s record in years. Clinical characteristics were extracted in the form of two digit diagnosis codes following the International Classification of Diseases v10 (ICD-10). Clinical features were extracted until the date of ATLAS enrollment for the enrolled sample, and for the entire duration of the record for non-ATLAS participants. Patients were required to have ICD-10 codes outside the Z-chapter (marking miscellaneous, non-specific clinical encounters).

### Univariate analyses

To quantify the univariate influence on ATLAS enrollment, we applied a univariate logistic regression model on each of the features. These models were implemented using a generalized linear model with a binomial family in the *statsmodels* package in Python. Multi-level and categorical features were one-hot encoded.

To assess the robustness of discovered associations over time and across clinical encounter types, we performed sensitivity analysis of 12 generated cohorts with varying parameters. We varied the minimum number of clinical contacts (>=1 or >=2) and types of clinical encounters (all clinical encounters or those specific to the following categories: Appointment, Office Visit, Hospital Encounter, and Lab visit). Since ATLAS recruitment efforts changed over time, we split the four cohorts using each year between 2021 and 2023 as a threshold and concluded with 12 datasets.

Year thresholds were chosen based on known modifications to recruitment strategy with each year. Specifically, the program was launched in specific UCLA clinics and gradually expanded to include other clinical departments over time. Year 2020 marked the beginning of dissemination of universal consent via patient portals, but also marked suspensions or modifications to the ATLAS enrollment protocol through 2021 as a result of restrictions due to COVID-19.

### Multivariate random forest models

Models were constructed using a 1:1 downsampling approach, where 104,516 individuals not enrolled in ATLAS were randomly chosen for comparison against the 104,516 ATLAS enrolled individuals. This ensured calibrated model probabilities, and in turn, model-based weight estimates for weighted analyses.

We split our dataset into 10 equally sized folds, balancing the proportion of ATLAS-enrolled and -unenrolled samples. We trained a random forest (RF) model on nine folds and evaluated the model on the remaining tenth fold, where each fold was used for evaluation once. We compute model performance using the area under the receiver operating characteristic (AUROC) for each fold.

Further, in order to avoid recovery of overfitted enrollment probabilities for samples in a given training data split, enrollment probabilities were recovered for each individual from the model where that sample was in the held-out evaluation fold.

In order to decrease the feature set for predicting enrollment status effectively, we utilize a recursive feature elimination strategy, compressing the model specification to 5, 10, and 15 features respectively, while preserving model performance.

Models were implemented using the *sklearn*^*38*^ library in Python3.

Feature importances in the form of Shapley values were computed using the TreeExplainer functionality in the *shap*^*39,40*^ package.

### Construction of probability weights

Enrollment probabilities for the ATLAS sample were extracted from the smallest RF model and transformed into weights for downstream models using the following formula, analogously to Schoeler, et al. (2023)^22^:

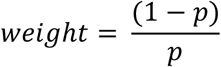

The behavior of this transformation can be best observed at its extremes, where an enrollment probability of 100% corresponds to a weight of 0, while an enrollment probability close to 0 corresponds to a large weight.

These probability weights were normalized by dividing by the mean of the overall weight distribution, ensuring that the sum of all probability weights equaled the available sample size.

### Polygenic scoring

We generate polygenic scores (PGS) for two traits: Major depressive disorder (MDD) and BMI. BMI scores (PGS ID: PGS002677) were generated from weights from the PGS catalog^32,41^. The MDD score was generated using the most recent meta-analysis^31^ and generated PGS using SBayesR^42^. PGS were residualized on 10 genetic PCs in the ATLAS cohort and z-score standardized.

### Replication of variant-level associations with select phenotypes

The phenotype-genotype reference map *pgrm*^*29*^ catalogs genetic associations spanning over 500 publications for replication studies in biobanks. We assess the replicability of findings in ATLAS under an unweighted and weighted framework using phenotypes encoded as phecodes. Using PLINK1.9^43^, we extract 4868 SNPs that intersect available ATLAS genotypes and span 5411 phecode associations from *pgrm*. We filtered phecodes that had low prevalence in ATLAS (<10%), retaining 1879 associations. For each association, we apply a linear model to associate genotype at the SNP to phecode endorsement in the ATLAS cohort, adjusting for the top 10 genetic PCs and sex. We also apply an analogous model, where the linear model was re-weighted with RF probability derived weights. Associations between SNPs and phecodes for each modeling scheme were compared against those reported in *pgrm*.

Associations were considered replicated for those with consistent direction with the *pgrm* catalog and using a nominal p-value threshold of 0.05, as the objective of this task was not discovery, but rather recovery of previously discovered associations. Z-tests of proportions were used to compare the proportion of replicated associations between model schemes.

### PGS analyses and phenome-wide association studies

We retrieved phecodes^44,45^ (v1.2) for all individuals for whom we have genetic data through the ATLAS biobank, mapping ICD-10 codes to existing phecode ontologies. For this analysis, phecodes were truncated at the level of 3 digit parent codes, reducing our available number of phecodes from 1841 to 560. Further, 5 phecodes were dropped due to missingness in the dataset, and 14 phecodes were dropped due to missing phecode metadata, leaving us with 541 phecodes. We performed a phenome-wide scan for each of the two PGS, using unweighted and weighted linear models to evaluate association of phecodes with each of the two PGS. Models were adjusted by self-identified sex. Significance was assessed using Bonferroni corrected correction at alpha level of 0.05 (p < 9.24e–5) to account for multiple testing.

To assess enrichment of associations corroborated by the weighting scheme compared to the unweighted scheme for clinically relevant phecode groups (see Supplementary Table 7), we utilized Fisher’s exact test, cross-tabulating membership in a phecode category and whether the association was shared by both unweighted and weighted models.. Significance was assessed using a cutoff of p < 0.05/17, which marks Bonferroni correction for the number of phecode categories tested.

To explicitly account for the 5 features used to generate inverse-probability weights in our unweighted associations, we also ran unweighted PGS-PheWAS associations including the top 5 features from the reduced RF models and compared associations. Feature level statistics were extracted for the 5 reduced RF model features. We further assessed whether associations that were significant in only the unweighted PheWAS as well as significant associations from both unweighted and weighted models remained significant in the feature-adjusted unweighted models.

In line with previous work^22,46–48^, we apply a linear model directly to our data. While this makes the assumption of a Gaussian distribution over a continuous outcome, it allows for a well defined convergence criterion in weighted regressions. While effect size point estimates are not identical to a linear regression with a binomial family, the direction of effect and trends in significance are consistent.

## Discussion

Biobank studies are prone to inclusion biases. Quantifying their extent, impact on genetic analyses, and developing analytic mitigation strategies, is crucial for making accurate inferences from these studies. In this work, we assessed inclusion bias in a clinically-linked biobank study, comparing the UCLA ATLAS biobank to the background UCLA Health cohort as a case study. We observed demographic and healthcare utilization features that distinguished the ATLAS subsample from the UCLA health system and, taken together, can be used to predict enrollment. Our findings suggest that correcting for bias through inverse probability adjustment increases power for genetic analyses (we observed an increase of 54% in replication rate of known GWAS variants) and may be used to reduce false positive associations.

Factors found to be associated with enrollment (such as XX, XX, XX) in univariate associations were stable across various sensitivity analyses accounting for primary care status, subsets of hospital visit types, and data horizons reflecting recruitment periods. Estimates of probabilities of enrollment from both the complete and reduced random forest models were driven, in large part, by demographic characteristics, corroborating strong associations observed in the univariate setting. While these demographic features are often correlated with clinical features, we observe a stronger association with frequency of healthcare utilization than any particular diagnostic markers. Further contextualizing these patterns of healthcare utilization according to types of clinical encounters (inpatient/outpatient/emergency) and level-of-care may elucidate relationships with enrollment patterns.

The increase in significantly replicated associations from the phenotype-genotype reference map we observe under the weighting scheme suggests increased power for discovery when adjusting for inclusion biases. Notably, weighting recovered functionally relevant associations, highlighting the *PPARG* and *CELSR2* genes along with genomic regions that have been linked with phenotypes such as diabetes and coronary atherosclerosis, indicating that adjustment for inclusion biases can aid in recovering biologically meaningful associations.

We observed a higher number of significant associations under unweighted analysis than weighted in both the MDD and BMI PGS-PheWAS settings. However, when categorizing phecodes within the relevant chapters (e.g. mental disorders for associations with MDD PGS, circulatory system for associations with BMI PGS) associations were more often significant in the weighted model than in unweighted model settings alone. These observations are consistent with our expectation that while relevant associations are faithfully recovered after employing our weighting scheme, spurious associations may be introduced through participation bias; however, this may also partially reflect the lower effective sample size in inverse probability weighted models^49^.

The heterogeneous effect of including important model features directly into the PheWAS analyses, reinforced our decision to use aggregate enrollment probability-derived weights, as opposed to the direct inclusion of model features as covariates. Instead of unadjusted covariates, these weights are directly informed by the enrollment outcome and provide a flexible framework for adjustments to downstream modeling tasks. While accounting for bias in a case-by-case manner specific to a given phenotype-genetic value pair might be useful, it entails significant analysis and cannot be uniformly applied in phenome-wide study contexts. Taken together, in addition to discovery efforts, we propose a primary use of this weighting scheme in PheWAS is to flag potential false positive associations that would emerge in unweighted analyses. We also note that the framework used here will not be suitable for rare variant studies as reweighting variants with low frequency may dramatically skew associations.

### Directions for future work

Our work highlights several directions for future work. First, we tested only a single and simple re-weighting scheme. Future work will involve the exploration of other strategies to account for such bias, including variations that enable rare variant associations.

Second, UCLA ATLAS is one in a growing collection of health-system linked biobanks. We hypothesize that the healthcare utilization and demographic characteristics that predict enrollment in ATLAS will be shared in other biobank settings. Additional studies in other such biobanks are vital to improve our understanding of inclusion biases and its effects. Moreover, while our results showed stability of feature importances over time, our study uses a release of genetic data from ATLAS including 54,770 samples. As the ATLAS Initiative continues to grow towards its initial target of 150,000 participants^25^, enrollment probabilities and weights will need to be updated in the future.

Third, comparing biobank participants to other patients served by the same health system has the advantage of the availability of rich data without the need for data harmonization. This choice of reference, however, is important. In future work, in line with previous work^22^, participation can be compared to the local population (e.g. city, county, or national population) using census measurements as a reference. These comparisons will be restricted to features available in both the biobank and reference data, requiring extensive efforts for feature alignment. ATLAS is one of the most diverse biobanks in terms of genetic ancestry^2^, serving a diverse metropolitan area. However, in this work, we did not attempt to compare ATLAS to the Los Angeles community as a whole.

Finally, it is important to note that inclusion bias consists of both recruitment and participation biases, where recruitment bias pertains to offering enrollment to patients and participation bias pertains to patients accepting the enrollment offer. Observationally, these effects are difficult to separate without additional information. Future work will involve the comparison of individuals who opted into the biobank to those who explicitly declined enrollment to disentangle biases induced due to recruitment and participant opt-in decisions. Insights from these analyses can potentially inform future recruitment strategies.

Taken together, in this work we emphasize the importance of considering inclusion biases in volunteer cohort study settings, with a focus on contextualizing discoveries that impact the clinical care setting. Accounting for factors related to enrollment can inform recruitment protocols, while also allowing for explicit post-hoc model adjustment in genetic investigations. With the growing utilization of genetic results in clinical contexts for screening^50^ and return of actionable results, inferences drawn from volunteer populations ought to be contextualized and calibrated to represent the background non-research participant population.

## Data Availability

Descriptive and summary statistics are contained within the manuscript and supplementary information. All individual-level patient records and genetic data are not available due to patient confidentiality and security concerns.

## Acknowledgements

The research reported here was supported by T15 LM013976 (to AP), R00 MH116115 (to LMOL), R01 MH137219 (to LMOL), U01 MH125042 (to LMOL), and U01 HG011715 (to BP).

We gratefully acknowledge the resources provided by the Institute for Precision Health (IPH) and participating UCLA ATLAS Community Health Initiative patients. The UCLA ATLAS Community Health Initiative in collaboration with UCLA ATLAS Precision Health Biobank, is a program of IPH, which directs and supports the biobanking and genotyping of biospecimen samples from participating UCLA patients in collaboration with the David Geffen School of Medicine, UCLA CTSI, and UCLA Health. Additionally, we greatly acknowledge Daniel H. Geschwind, Clara Lajonchere, and Maryam Ariannejad for their insightful feedback.

## Notes

### Competing Interest Statement

The authors have declared no competing interest.

### Author Declarations

Patient Recruitment and Sample Collection for Precision Health Activities at UCLA is an approved study by the UCLA Institutional Review Board (UCLA IRB). IRB#17-001013.

