## Supplementary material for "Inclusion bias affects common variant discovery and replication in a health-system linked biobank": Figures and Supplementary Figures

**Main Figures & Tables**

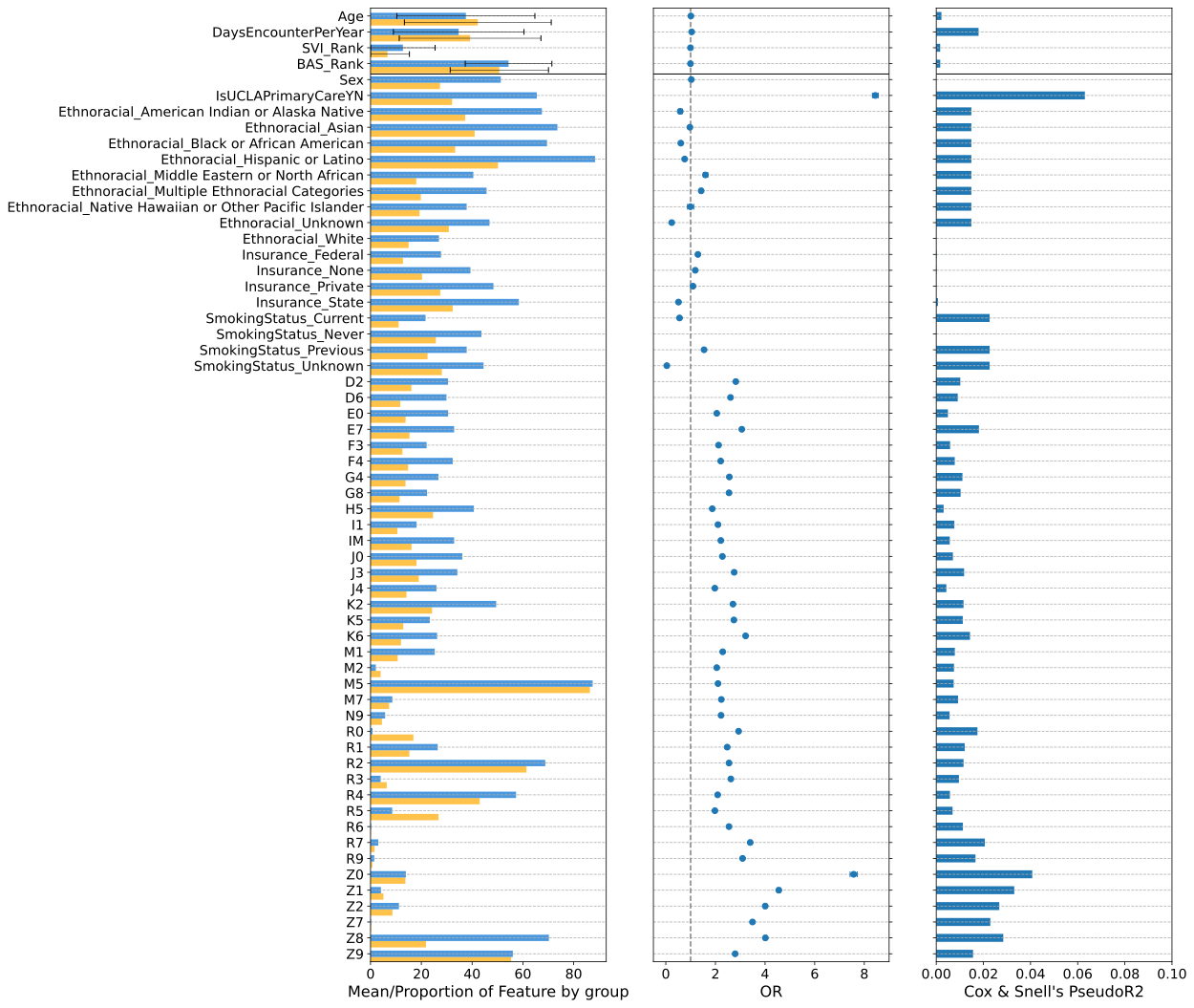

**Figure 1:** Cohort characteristics of the UCLA health sample and ATLAS subsample. (Left) Feature level distributions stratified by ATLAS enrollment (yellow = not enrolled in ATLAS, blue = enrolled in ATLAS). For quantitative variables, feature means are depicted, and for categorical features, proportion of individuals in both groups in the feature group are depicted. For diagnostic chapters, any diagnoses in the chapter is sufficient. (Middle/right) Results from univariate associations with ATLAS enrollment Odds ratio (middle) and Cox & Snell’s pseudo-R2 (right).

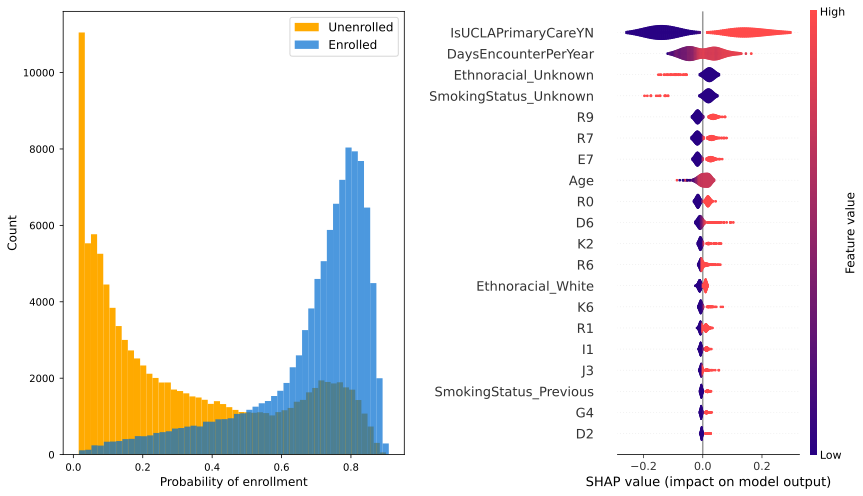

**Figure 2:** Probability distributions stratified by enrollment and feature characteristics from a multivariate random forest model classifier of ATLAS enrollment. (Left) Predicted probability distributions stratified by true ATLAS enrollment status (yellow = not enrolled in ATLAS, blue = enrolled in ATLAS) show strong separation between the classes. (Right) Beeswarm plot of Shapley values on a subset of 100 individuals reveal healthcare utilization patterns and select ICD-10 diagnoses as important predictors of ATLAS enrollment.

|  | **Unweighted** | **Weighted** | **Shared** | **Unweighted only** | **Weighted only** |
| --- | --- | --- | --- | --- | --- |
| # replicated associations (p < 0.05) | 606 | 724 | 389 | 217 | 335 |
| # phecodes with replicated associations (p < 0.05) | 21 | 23 | 20 | 19 | 20 |

**Table 1:** Counts and proportions of replicated associations from pgrm in the ATLAS sample. Associations were considered replicated at a significance level of p < 0.05 and when direction of effect was consistent with pgrm.

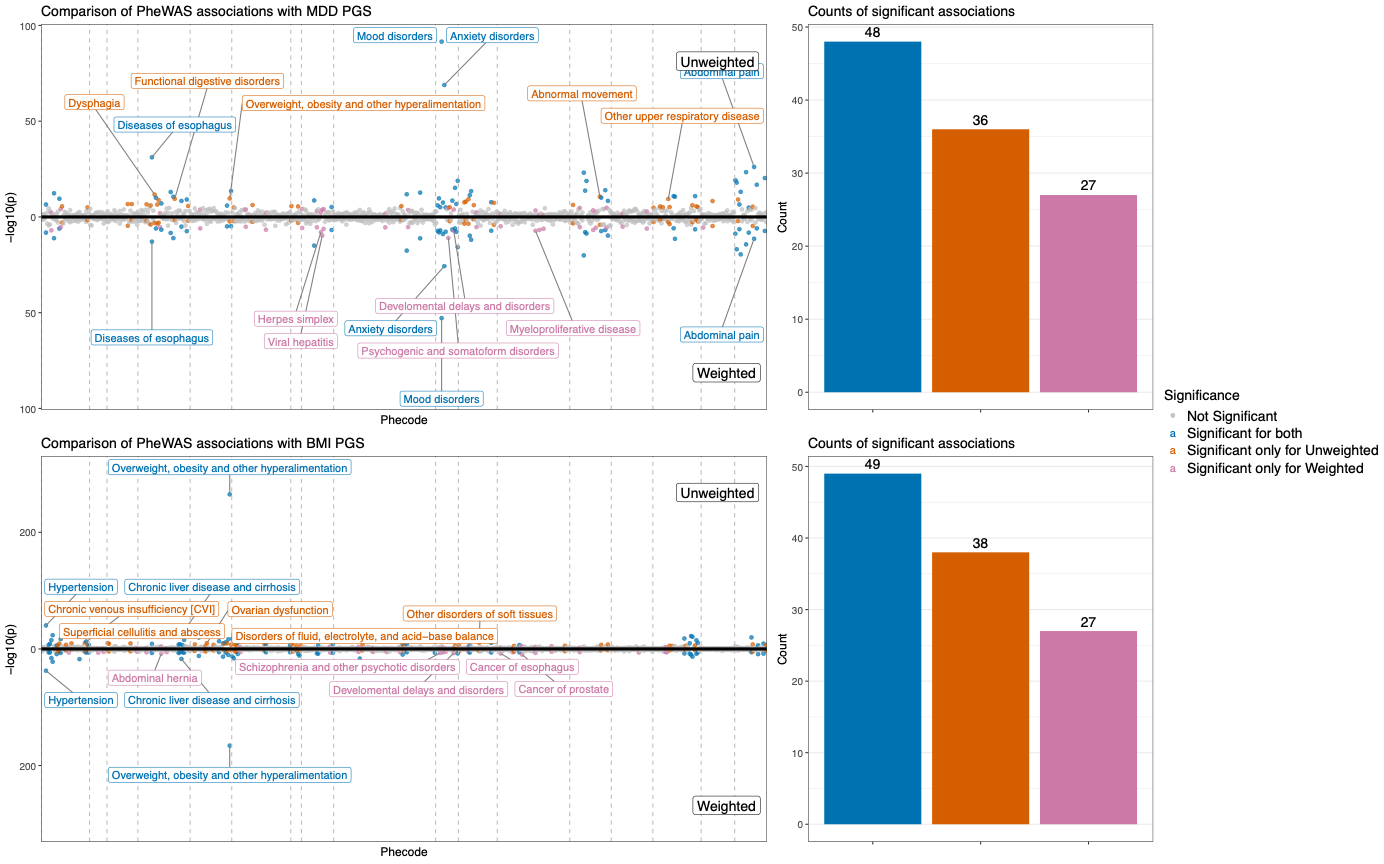

**Figure 3:** Comparison of weighting schemes in phenome-wide associations with PGS. Miami plots for PheWAS on PGS for MDD and BMI (top: unweighted, bottom: weighted) show shared and unique associations under the unweighted and weighted models.

**Supplementary Figures**

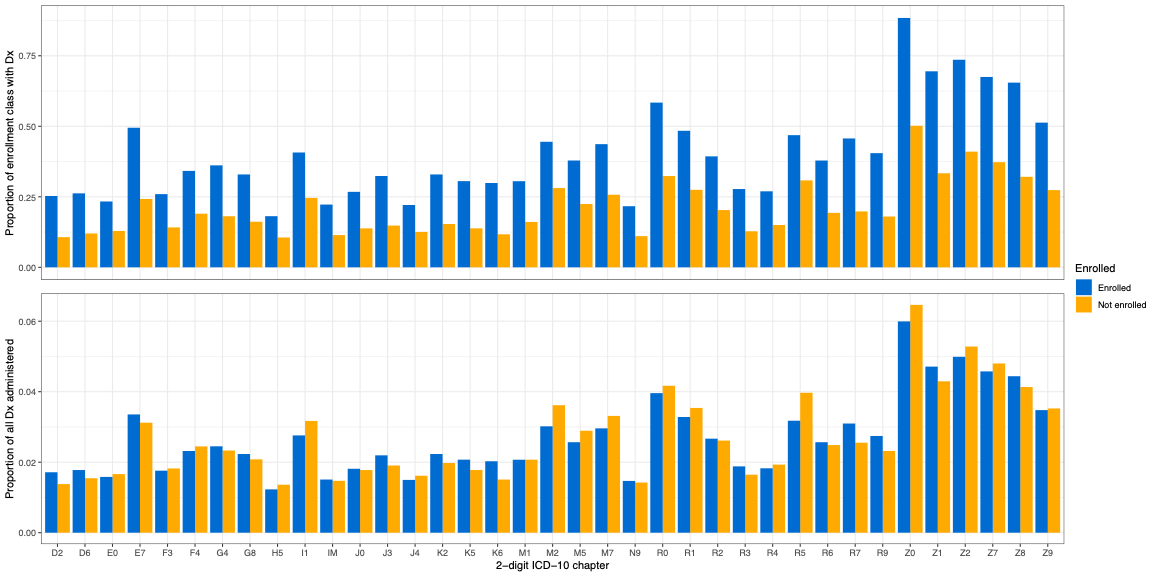

**Supplementary Figure 1:** Comparison of diagnostic burden across enrollment groups. Proportion of individuals in each enrollment category with a given 2-digit ICD-10 diagnosis (top) and proportion of all diagnoses administered in a given enrollment category comprised by each 2-digit ICD-10 code (bottom)

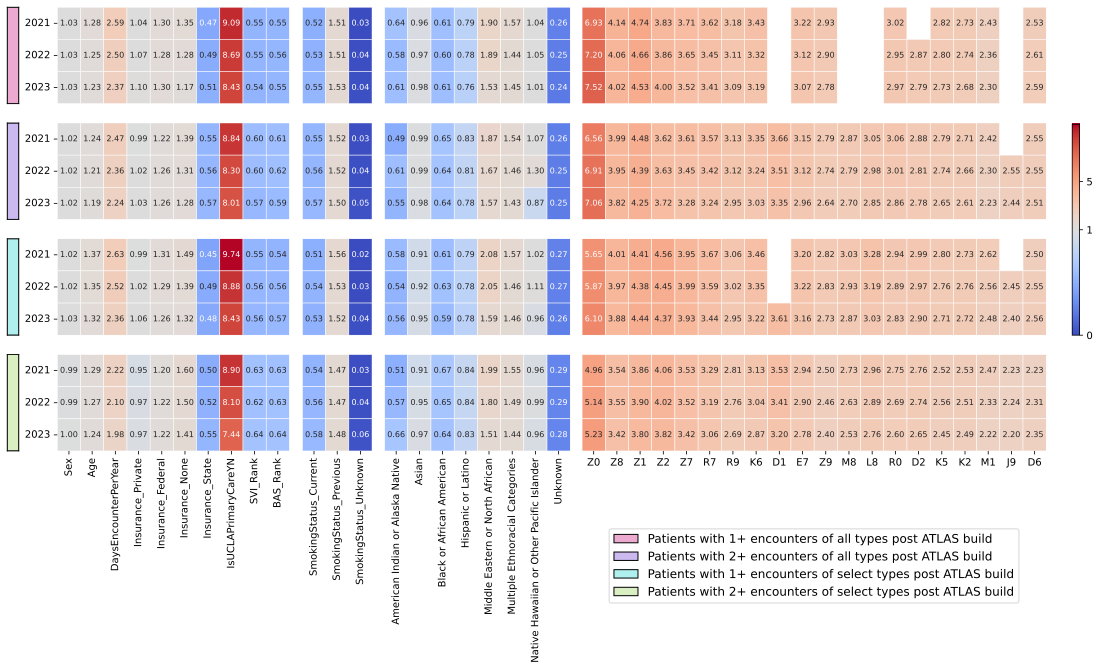

**Supplementary Figure 2:** Comparison of feature level effects on enrollment across various encounter types and recruitment period data horizons

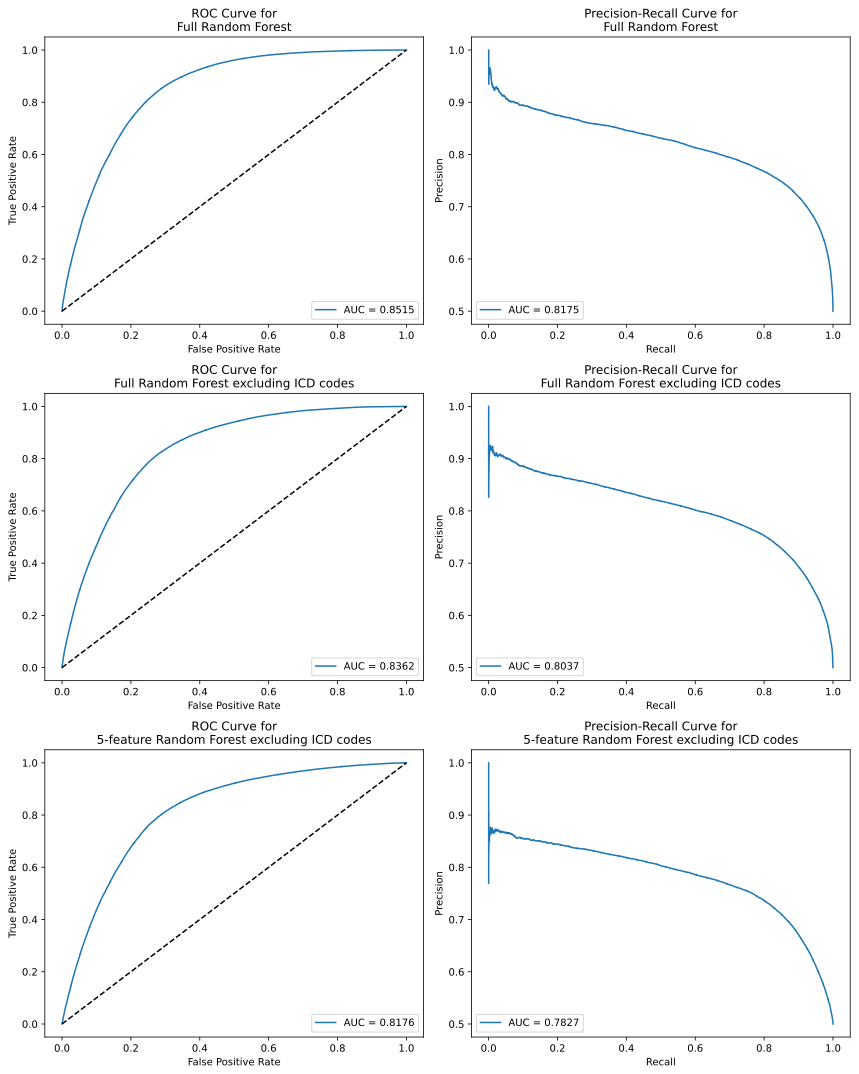
**Supplementary Figure 3:** Random forest model performance across model settings (full model, full model excluding ICD-10 codes, 5-feature recursive feature elimination)

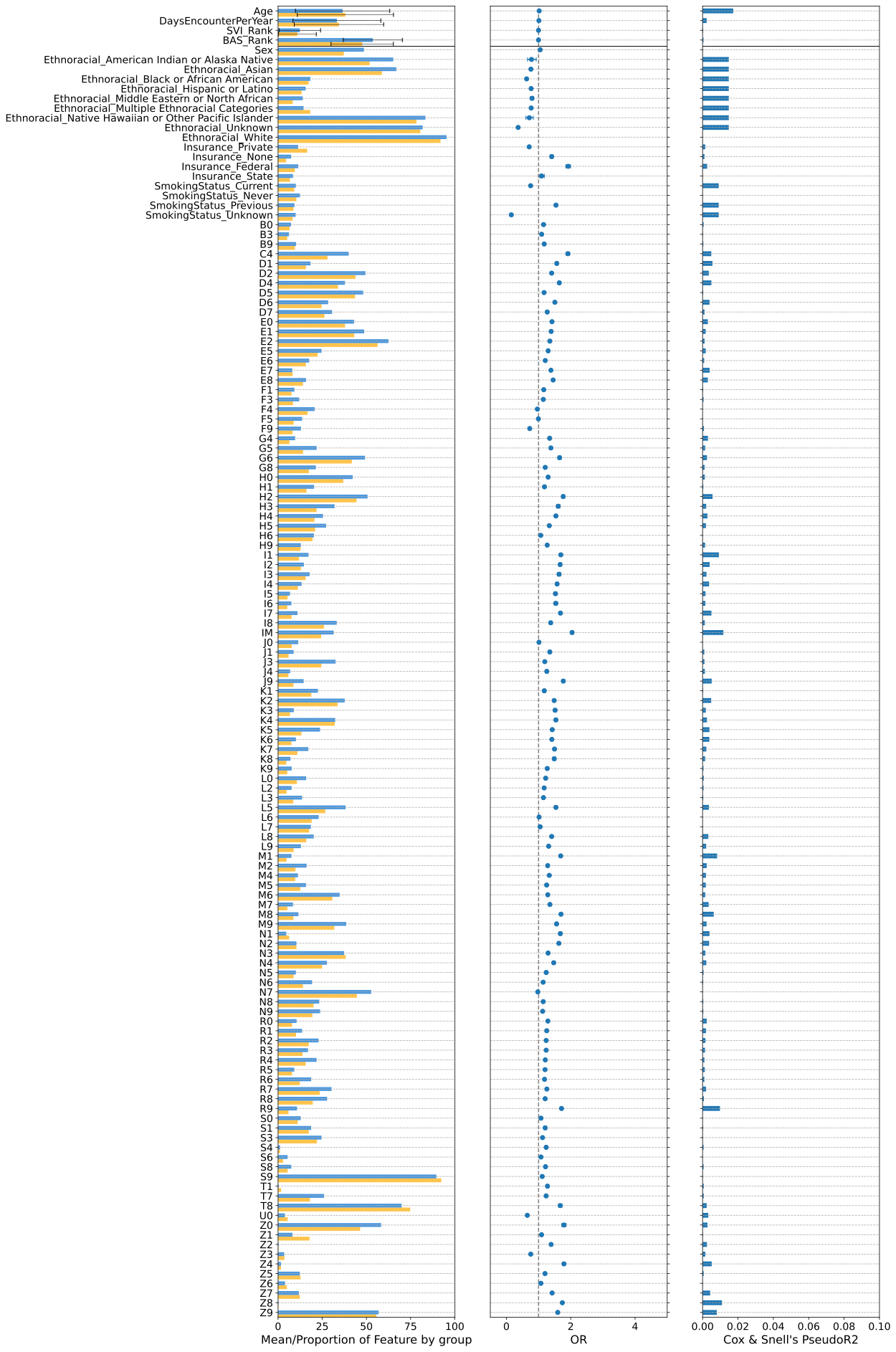

**Supplementary Figure 4:** Feature distributions and univariate association statistics for ATLAS enrollment in a subset of individuals receiving primary care at UCLA

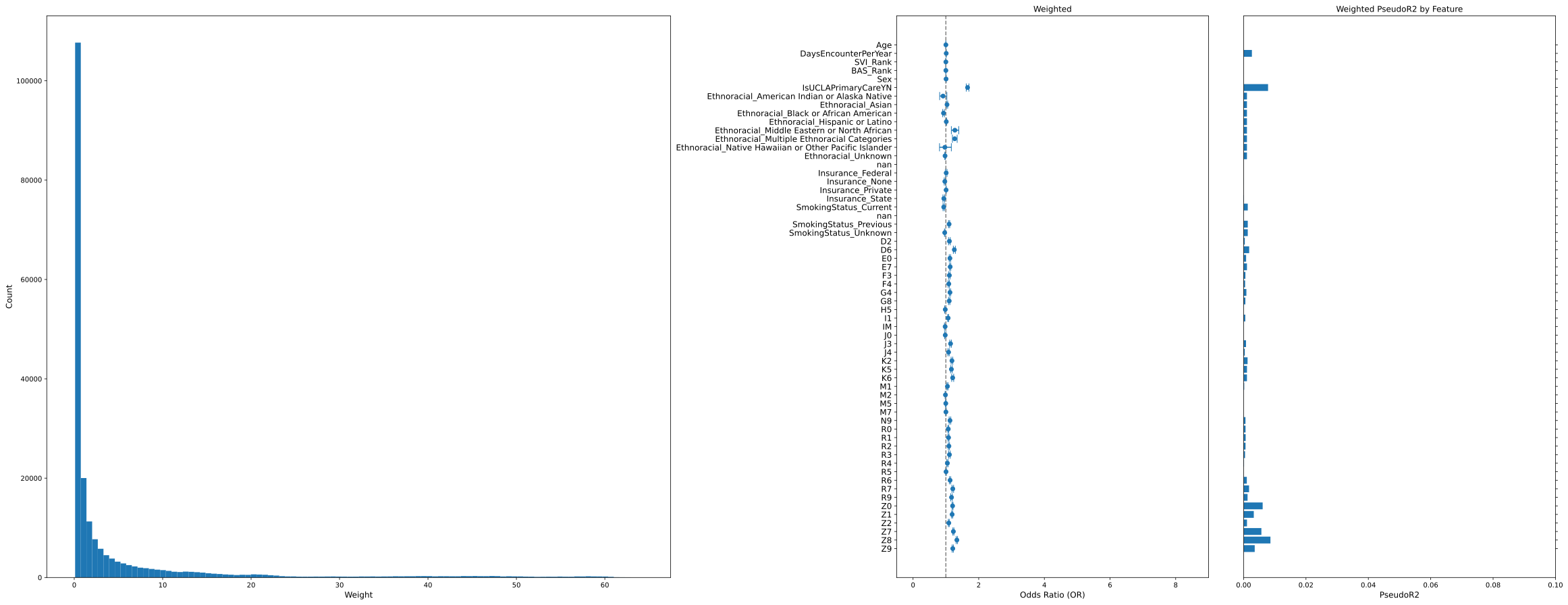

**Supplementary Figure 5:** Distribution of weights (left) and weight-adjusted univariate associations (right) after applying inverse probability weighting scheme using random forest model estimates

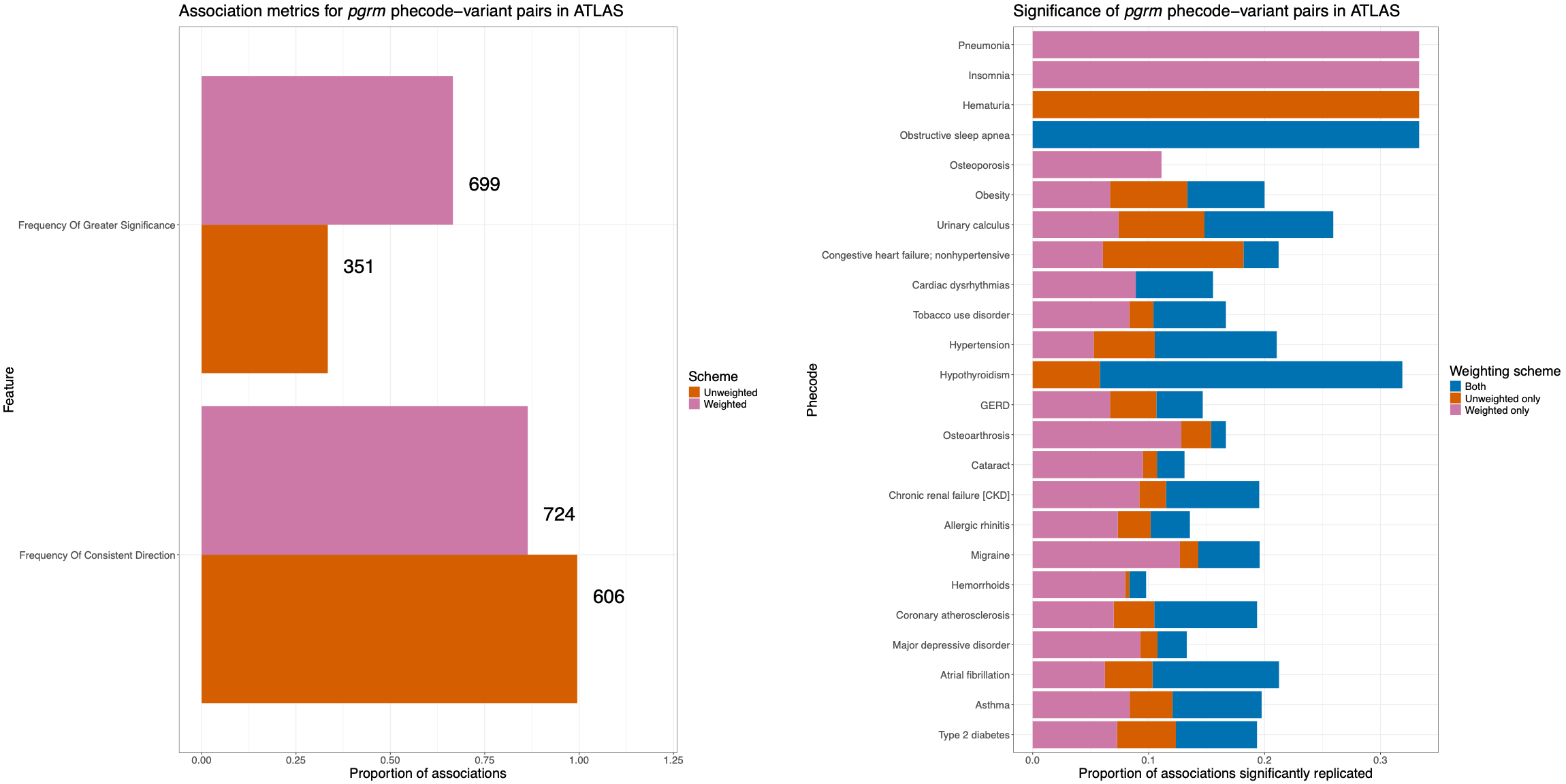

**Supplementary Figure 6:** Breakdown of replication of pgrm phecode-variant pairs based on frequency of greater significance and consistent direction (left) and phecode category (right)

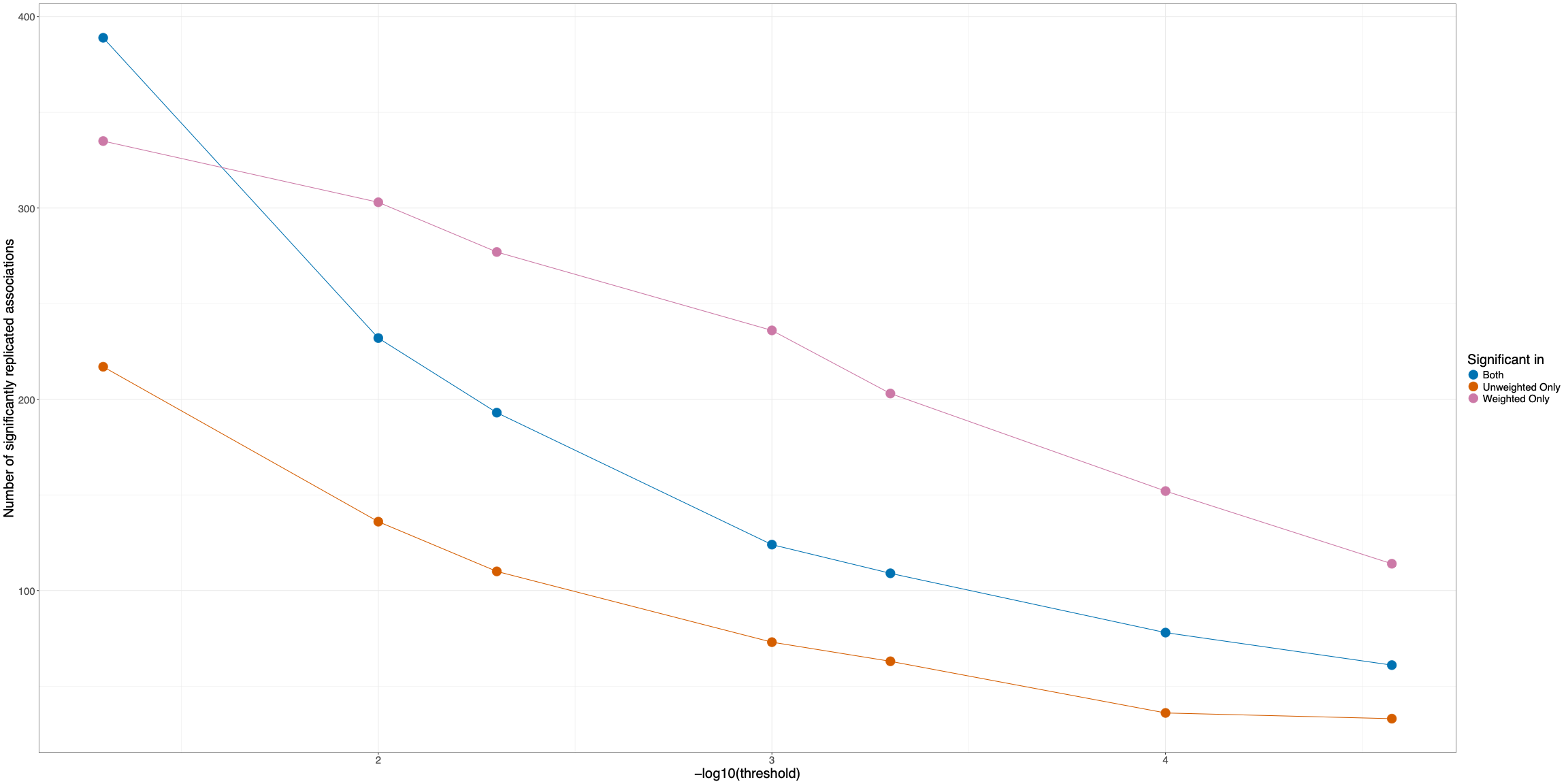

**Supplementary Figure 7:** Number of significantly replicated pgrm associations at varying significance thresholds (p < 0.05, 0.01, 0.005, 0.001, 5e-4, 1e-4, Bonferroni corrected 2.66e-5)

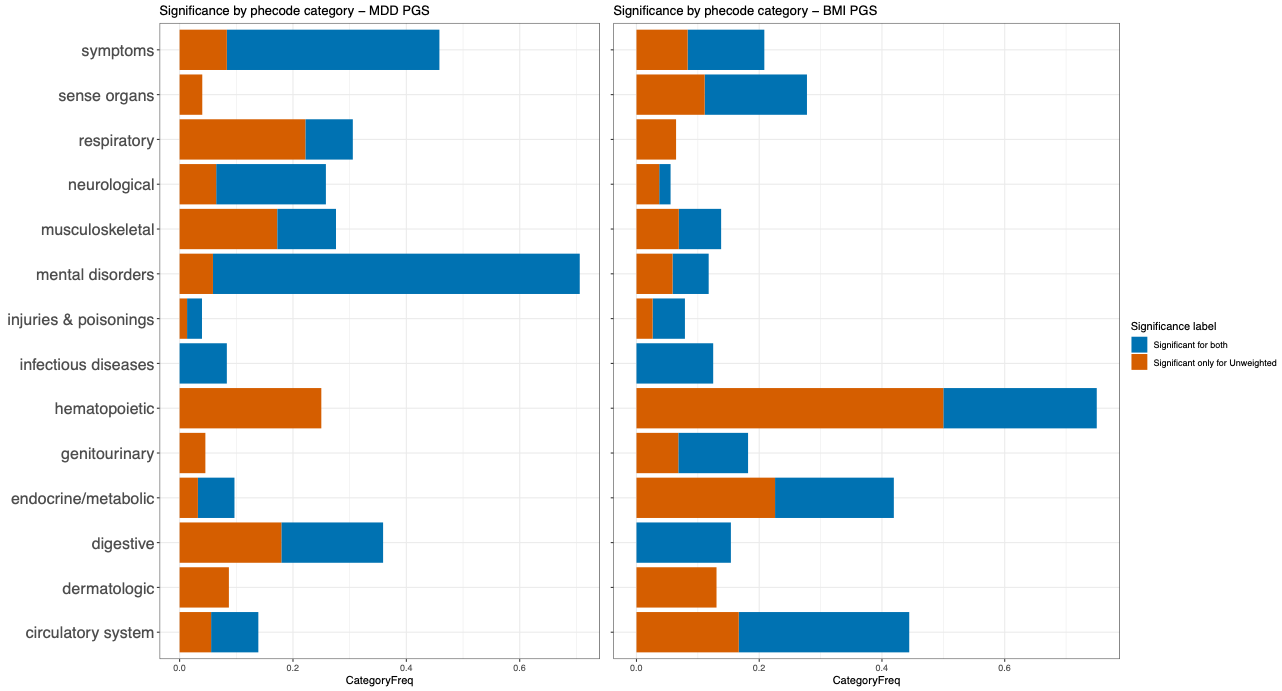

**Supplementary Figure 8:** Breakdown by phecode category of significant PheWAS associations with MDD PGS (left) and BMI PGS (right)

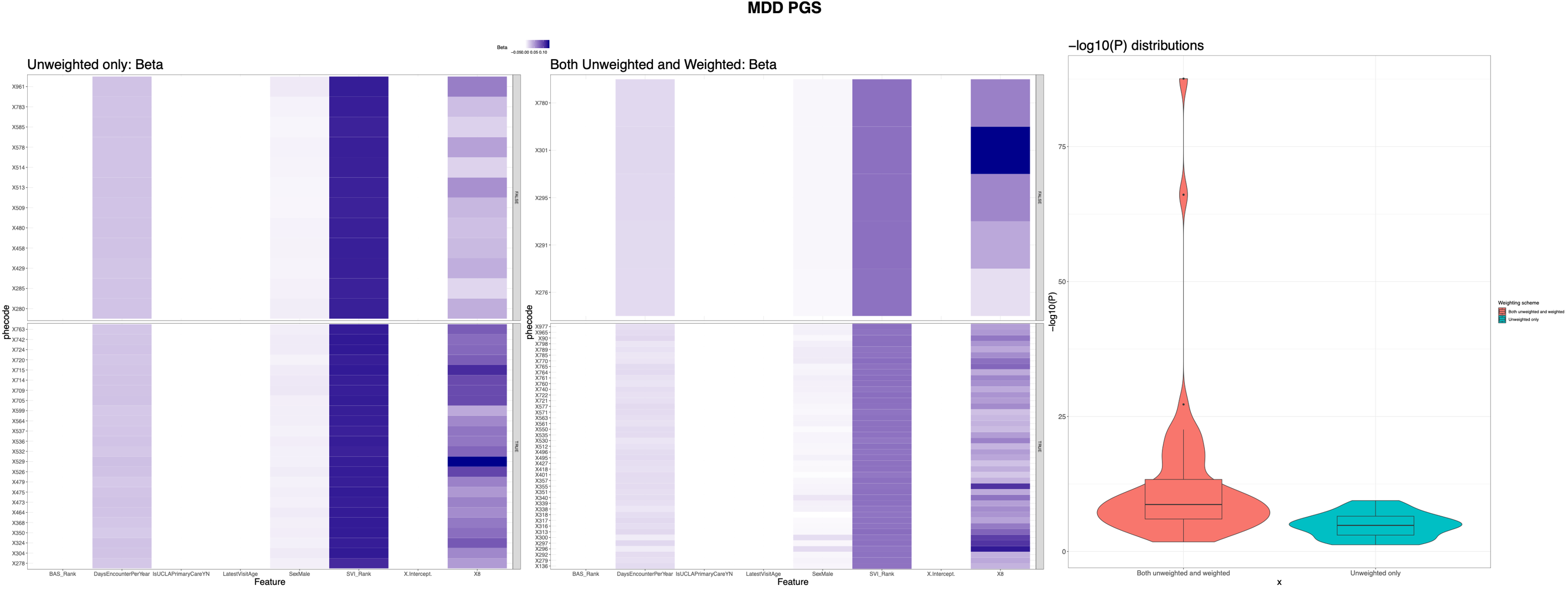

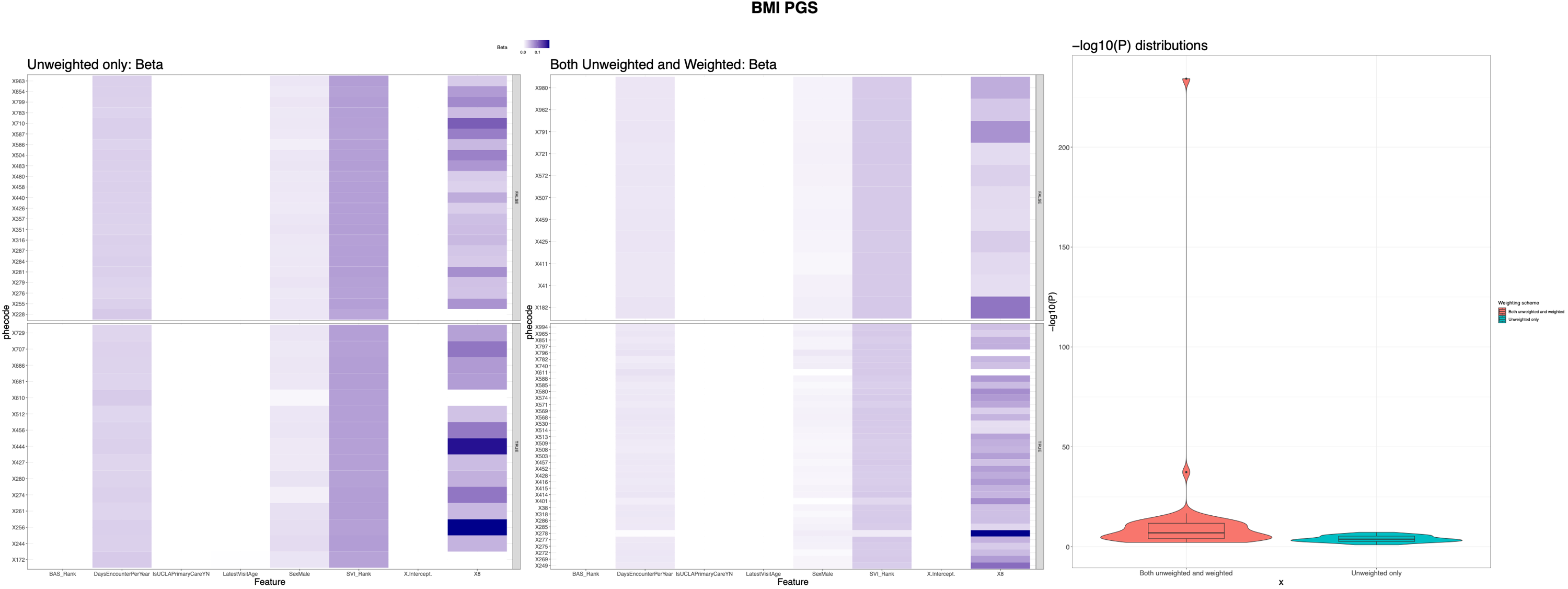

**Supplementary Figure 9:** PheWAS associations with MDD PGS (top) and BMI PGS (bottom) when directly adjusting for reduced random-forest model features. Phecodes are grouped by significance in unweighted and weighted PheWAS comparing significant associations in only unweighted settings (left) to significant associations under both weighting schema (middle). Log(P) distributions stratified by significance are shown on the right.
